# Multivariate patterns linking brain microstructure to temperament and behavior in adolescent eating disorders

**DOI:** 10.1101/2024.11.24.24317857

**Authors:** Carolina Makowski, Golia Shafiei, Megan Martinho, Donald J. Hagler, Diliana Pecheva, Anders M. Dale, Christine Fennema-Notestine, Amanda Bischoff-Grethe, Christina E. Wierenga

**Author notes:** Authors share senior authorship (Amanda Bischoff-Grethe, Christina E. Wierenga). **Correspondence to:** C.M.

## Abstract

Eating disorders (EDs) are multifaceted psychiatric disorders characterized by varying behaviors, traits, and cognitive profiles thought to drive symptom heterogeneity and severity. Non-invasive neuroimaging methods have been critical to elucidate the neurobiological circuitry involved in ED-related behaviors, but often focused on a limited set of regions of interest and/or symptoms. The current study harnesses multivariate methods to map microstructural and morphometric patterns across the entire brain to multiple domains of behavior and symptomatology in patients. Diffusion-weighted images, modeled with restriction spectrum imaging, were analyzed for 91 adolescent patients with an ED and 48 healthy controls. Partial least squares analysis was applied to map 38 behavioral measures (encompassing cognition, temperament, and ED symptoms) to restricted diffusion in white matter tracts and subcortical structures across 65 regions of interest. The first significant latent variable explained 46.9% of the covariance between microstructure and behavior. This latent variable retained a significant brain-behavior correlation in held-out data, where an ‘undercontrolled’ behavioral profile (e.g., higher emotional dysregulation, novelty seeking; lower effortful control and interoceptive awareness) was linked to increased restricted diffusion across white matter tracts, particularly those joining frontal, limbic, and thalamic regions. Individually-derived brain and behavior scores for this latent variable were higher in patients with binge-purge symptoms, compared to those with only restrictive eating symptoms. Findings demonstrate the value of applying multivariate modeling to the array of brain-behavior relationships inherent to the clinical presentation of EDs, and their relevance for providing a neurobiologically-informed model for future clinical subtyping and prediction efforts.

## INTRODUCTION

Eating disorders (EDs), including anorexia nervosa (AN) and bulimia nervosa (BN), comprise serious psychiatric conditions characterized by dysregulated eating behaviors and significant concerns around weight and body shape. The onset of EDs typically occurs during adolescence [1], a critical time period for cognitive and brain development. There has been a growing appreciation for the role of neurobiology in the etiology and maintenance of EDs [2–5], yet clear brain-based biomarkers remain undefined.

The majority of non-invasive neuroimaging studies in EDs have turned to diagnostic ‘case-control’ frameworks, but have failed to find robust and reproducible neural markers that are either predictive of EDs or prognostic indicators of treatment outcomes. Case-control designs operate under the misleading assumptions that the group mean is representative of individual patients, and that diagnostic categories will map onto biological mechanisms [9]. In reality, diagnostic categories pool across heterogeneous clinical and behavioral presentations of a given disorder, clouding efforts to uncover meaningful biological differences between groups that are needed to push our neurobiological understanding of EDs forward. Understanding individual patient differences and how they map onto neuroimaging-derived measures also requires methods that move beyond assigning a symptom or behavior to one or a limited set of brain regions [10, 11]. The current work takes a departure from conventional diagnostic and univariate study designs, and instead adopts a data-driven approach to parse apart the heterogeneity of clinical presentations of EDs and to magnify associations with an array of neuroanatomical measures.

Acknowledging and incorporating the array of behavioral and cognitive factors contributing to ED symptomatology into study designs is key in understanding the factors that may contribute to chronic and non-remitting illness courses in many patients [6, 12, 13]. For instance, specific profiles of temperament (e.g., harm avoidant, perfectionistic, inhibited behavior) and emotional dysregulation have been shown to be present before ED onset and even persist after recovery [14]. Under-vs. over-controlled ED subtypes, which tend to characterize patients with binge-purge and restricting symptoms, respectively, have been described to more succinctly capture personality profiles of patients with EDs that are independent of diagnostic classification [15–17]. The genetic basis of the traits contributing to these personality profiles are, in part, expressed through alterations in neurotransmission and neurocircuitry [18–21], which in turn may predispose an individual to an ED later in life. Other behaviors and cognitions may reflect more transient states due to illness, such as body image concerns and severity of disordered eating behaviors. Behavioral predispositions to ED symptoms are also linked to internal sensations, such as interoceptive awareness [22], or cognitive performance that may improve with treatment, such as working memory and executive functioning [23–25]. Capturing the interplay between various personality and symptom constructs in EDs with neuroimaging data may be critical to parse apart the heterogeneity of EDs in a neurobiologically-informed manner.

Many of the above-mentioned behaviors and symptoms have neurobiological underpinnings. However, it is unclear how these behavioral and clinical profiles collectively map onto brain structure in adolescent ED patients, which could be elucidated with multivariate methods that map onto features derived from non-invasive brain magnetic resonance imaging (MRI). The majority of neuroimaging studies in ED patients have focused on structural morphometry, such as cortical thickness and volumes [26]. There is strong evidence to suggest that alterations in these morphological measures are largely due to undernutrition and are reversed upon weight restoration [27–29]. There have been far fewer studies applying diffusion imaging to understand the neurobiology of EDs compared to structural MRI studies [26, 30]. However, diffusion imaging provides an important opportunity to understand how white matter tissue microstructure may predispose and/or contribute to maintenance of ED symptoms, given the ongoing development of white matter microstructure throughout adolescence. There is also evidence of sex differences in white matter development (e.g., earlier maturation in females compared to males) [31], suggesting microstructural markers may be important features to study to understand the higher prevalence rates of EDs in females [32]. Existing diffusion studies in adolescents with EDs have yielded discrepant findings, particularly with fractional anisotropy (FA; degree of oriented diffusion along tracts), with studies reporting both increases and decreases in FA across various tracts [33–35]. Some reports have failed to find significant case-control differences altogether [36, 37].

Compared to the commonly used diffusion tensor model, advances in diffusion-weighted imaging acquisitions can offer a more fine-grained interpretation of diffusion within intra- and extra-cellular tissue compartments, as well as more accurate modeling of crossing fibers and microstructure beyond major white matter tracts [38, 39]. One report found reduced neurite density in fibers joining the ventral tegmental area and nucleus accumbens in young patients with AN compared to controls [40]. However, the study was limited by its focus on two tracts of interest. One advanced diffusion model, restriction spectrum imaging (RSI) [38] offers additional neurobiological insight beyond the classic diffusion tensor model as it differentiates between intracellular and extracellular tissue compartments, and can also model diffusion emanating from multiple directions within a voxel. RSI has shown success in detecting meaningful associations with cancer [41, 42], neurological disorders [43–46], sex at birth [47], environmental exposures [48], and typical [39] and atypical brain development [49]. This model also allows for the investigation of microstructural properties of deeper subcortical structures [39, 50]. Subcortical structures are key regions of interest in ED-related brain circuitry, and have typically been studied with structural and functional imaging to better understand altered reward processing and learning [51–54]. RSI of brain microstructure has not yet been applied to the study of EDs, and could provide important mechanistic insights into ED pathophysiology.

The lack of consensus among the above-mentioned studies using diffusion-weighted imaging in EDs could be largely attributed to underpowered studies (N∼20-30 individuals per group), as well as adoption of univariate statistics to test case-control differences. Univariate methods have dominated the statistical landscape of not just ED neuroimaging research, but psychiatry at large. Univariate statistics can either lead to inflated rates of false positive findings that are hard to replicate in underpowered samples [55], or yield small effect sizes that do not survive correction for multiple comparisons [10, 56–58]. One of the above-mentioned studies reported null case-control findings with diffusion, but instead found that multivariate pattern detection was more fruitful in predicting ED status [37]. Shifting from classical univariate statistics to a multivariate framework may help us understand individual-level differences, which in turn, will be necessary for precision psychiatry efforts [10, 59].

The current study harnesses various methodological advances to parse heterogeneous clinical and behavioral profiles of EDs to patterns of brain microstructure in a well-powered transdiagnostic sample of adolescent girls. This includes: 1) a multivariate data-driven framework to map individual variability in brain-behavior patterns; 2) restriction spectrum imaging to map both white matter and subcortical tissue microstructure; 3) adolescent patients early in the course of illness, which could help mitigate effects of illness chronicity; and 4) a detailed survey of temperament, cognitive and other behavioral traits in both patients and controls, alongside ED symptoms in patients across two timepoints (baseline and one-year follow-up). Our objectives were three-fold. First, we aimed to map patterns of tissue microstructure across both white matter and subcortical structures to clinical and behavioral profiles in a well-powered and well-characterized sample of adolescent girls with an ED. Next, we derived individual-level scores to investigate the degree to which an individual patient’s data maps onto statistically robust patterns of brain and behavior. Finally, we tested the clinical utility of these derived scores, and investigated whether individual-level scores mapped onto clinically-derived diagnostic subtypes (e.g., binge-purge vs restrictive subtypes of EDs) and/or could be predictive of future clinical symptoms. Given the multivariate and data-driven nature of the proposed analysis, no *a priori* hypotheses were made about specific brain-behavior profiles that would emerge.

## METHODS

### Sample

Female adolescents with an ED, meeting criteria for a DSM-V restricting or binge-purge type eating disorder [60] [61], were recruited from the University of California, San Diego Eating Disorders Treatment and Research Program. ED diagnosis was determined by meeting *DSM-V* criteria through the Kiddie Schedule for Affective Disorders and Schizophrenia (KSADS-5) [62, 63], a semi-structured interview performed by a trained research assistant, under the supervision of a doctoral level psychologist. The ED-restricting subtype comprised patients with DSM-V diagnoses that included primarily restriction of food intake (Anorexia Nervosa-Restrictive subtype [AN-R], Atypical AN [AAN], and Avoidant Restrictive Food Intake Disorder [ARFID]); ED-binge purge subtype included patients with diagnoses characterized by binge and/or purge symptoms (Bulimia Nervosa [BN], AN-BN, AN with binge-purge symptoms [AN-BP], AN with purge symptoms [AN-P], Purging Disorder (PD), and Other Specified Feeding Disorder [OSFED] with PD features). HCs were recruited from the San Diego community and did not have any ED symptomatology or Axis I psychiatric disorder, as determined by the Eating Disorders Examination (EDE) and KSADS-5, respectively. See Supplement for additional exclusion criteria. All study procedures were approved by the University of California, San Diego’s Institutional Review Board (170664) and informed consent/assent was obtained prior to initiation of study procedures.

### Clinical and behavioral measures

We included measures from five domains: i) cognition, including cognitive flexibility, inhibition, abstract reasoning and verbal skills; ii) temperament, including reward and punishment sensitivity; iii) interoceptive awareness, including measures of body trust and attending to bodily sensations; iv) emotion recognition and regulation; and v) ED-related symptom severity, including cognitive concerns around weight, shape and eating, as well as physical symptoms such as restricting food intake, purging, and excessive exercise. See Table 1 and Supplementary Table 1 for a full list of included instruments and variables.

**Table 1.**
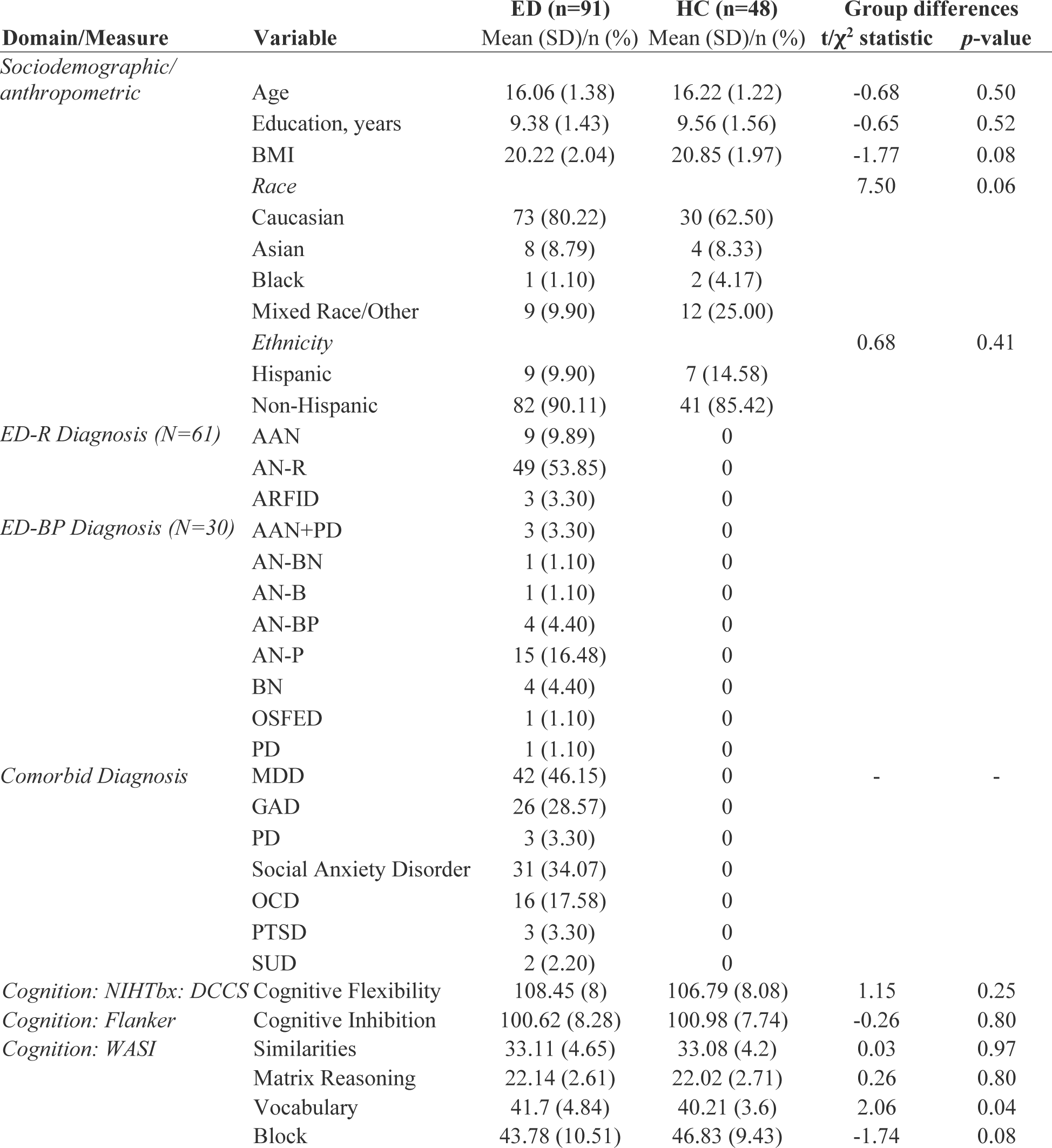
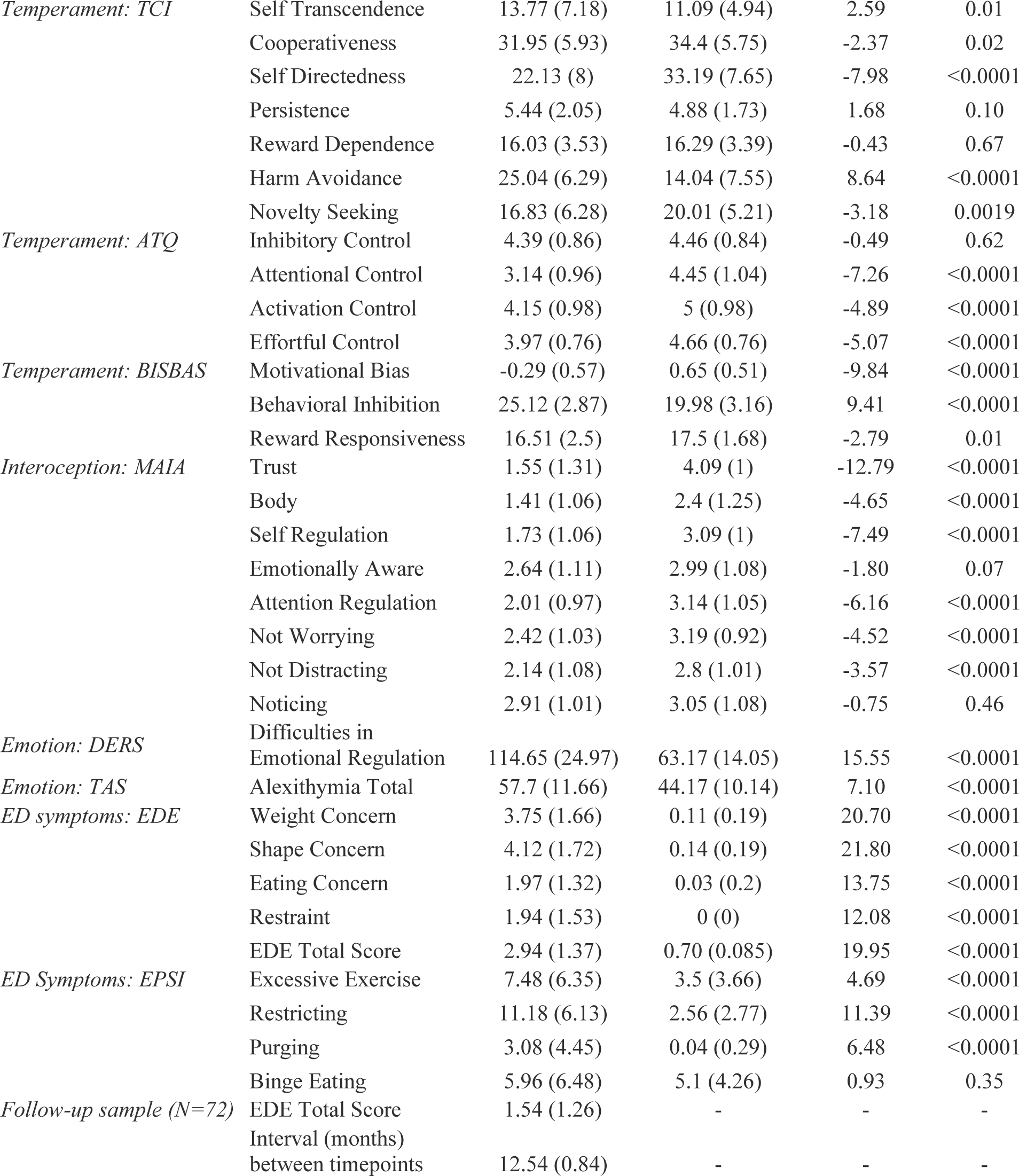
Demographic and behavioral descriptive statistics for included participants. Abbreviations: ED-R, Eating Disorder - restrictive subtype; ED-bp, Eating Disorder - binge purge subtype; HC, Healthy Control; DCCS NIHTbx: Dimensional Change Card Sort task from the National Institutes of Health Cognitive Toolbox [76]; Flanker: Flanker task [77]; WASI, Wechsler Abbreviated Scale of Intelligence (WASI) [78]; TCI, Temperament and Character Inventory (TCI) [79]; ATQ, The Adult Temperament Questionnaire [80]; BISBAS, Behavioral Inhibition and Behavioral Activation Scales [81]; MAIA, Multidimensional Assessment of Interoceptive Awareness [82]; EDE, Eating Disorders Examination [83, 84]; Eating Pathology Symptoms Inventory [85]; DERS, Difficulties in Emotion Regulation Scale [86]; TAS, Toronto Alexithymia Scale [87]; BMI, Body Mass Index. AAN, Atypical Anorexia Nervosa. AN, Anorexia Nervosa. ARFID, Avoidant/Restrictive Food Intake Disorder. AN-B, Anorexia Nervosa-Binge subtype. AN-BP, Anorexia Nervosa-Binge Purge subtype. AN-R, Anorexia Nervosa-Restricting subtype. BN, Bulimia Nervosa. OSFED, Other Specified Feeding Disorder. PD, Purging Disorder. MDD, Major Depressive Disorder. OCD, Obsessive Compulsive Disorder. PTSD, Post-Traumatic Stress Disorder. SUD, Substance Use Disorder.

### Image processing and derived variables

MRI Images were collected on a 3.0 T GE MR750 scanner equipped with quantum gradients providing echo planar capability, using a Nova Medical 32 channel head coil (maximum gradient strength: 50 mT/m, slew rate: 200 T/m/s). The following protocol was administered to acquire structural T1-weighted images and multi-shell diffusion: a three-plane localizer scan; a whole brain, sagittally acquired (0.8 mm slice thickness, FOV=256 mm) T1-weighted (MPRAGE PROMO, TE=3.656 ms, flip angle=8°, matrix=320, 2x in-plane acceleration) and separate T2-weighted (3D CUBE, 0.8 mm slice thickness, FOV= 256 mm, TE=60 ms, variable flip angle, matrix=320, 2x in-plane acceleration) sequence for alignment and morphometry; and two DTI scans (FOV=240 mm, slice thickness = 1.7 mm, matrix = 140 x 140, b=1500/3000 s/mm^2^; 102 diffusion directions; multiband factor=3), which were further used for restriction spectrum imaging (RSI) modeling.

Data were processed using the Adolescent Brain Cognitive Development (ABCD) Study pipeline, as described in [64]. See Supplement for additional details on image processing and derivation of region-of-interest inputs. Our main investigation focuses on restricted normalized directional (RND) diffusion across white matter tracts and subcortical structures. More details of the RSI model are included in Supplementary Materials and in previous publications [38, 39, 42]. Briefly, restricted diffusion describes water within intracellular spaces confined by cell membranes with a non-Gaussian pattern of displacement, where RND models diffusion emanating from multiple directions within a voxel. We measured RND in 35 white matter tracts from AtlasTrack [65] and 30 subcortical regions from the probabilistic subcortical segmentation (*aseg)* in Freesurfer [66]. See Supplementary Tables 2 and 3 for a full list of all regions of interest (ROIs).

### Partial Least Squares model and statistics

We used Partial Least Squares (PLS) to study the relationship between restricted diffusion patterns across white matter tissue and subcortical structures, and behavioral measures described above in our patient sample. PLS analysis is a multivariate statistical technique that identifies weighted patterns of variables in two sets of data that maximally covary with each other [67–70]. PLS is advantageous in that it does not rely on statistical independence of input variables and can handle collinearity between features. In the present analysis, one variable set corresponded to age-corrected restricted diffusion across 65 regions of interest, and the other to 38 behavioral (clinical, cognitive, temperament) measures. The two variable sets were normalized by z-scoring the variables across columns and correlated with each other across patients. The resulting correlation matrix was then subjected to singular value decomposition to identify latent clinical-anatomical dimensions (i.e., latent variables) that capture the maximal covariance between the two variable sets. See **Figure 1** for a schematic of our PLS analysis workflow.

**Figure 1.**
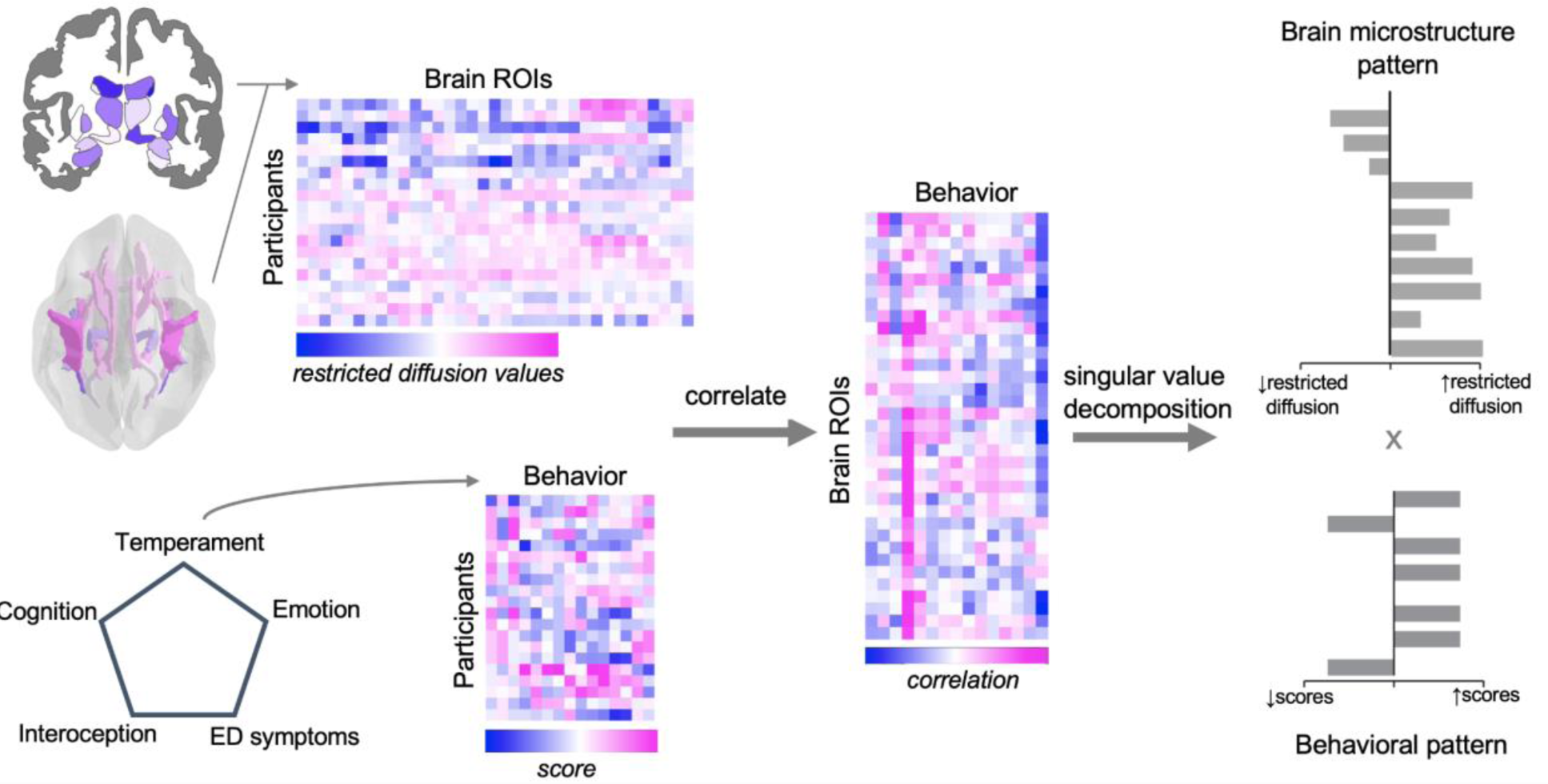
PLS analysis workflow.

Inference and validation of the statistical model were performed using nonparametric methods, as previously described [70–72]: i) statistical significance of overall patterns was assessed by permutation tests [73]; ii) feature (voxel, clinical-cognitive measure) importance was assessed by bootstrap resampling [74]; and iii) out-of-sample correlations between projected scores were assessed by cross-validation [71, 75]. More details of the analysis and inferential methods are described in supplementary material.

### Clinical applications of PLS analysis

To see whether behavioral patterns may relate to restricting vs binge-eating/purging subtypes of EDs, which are often described in clinical settings, brain and behavior scores from significant latent variables were compared between these two subtypes. To further explore the potential clinical utility of derived latent variables, we also investigated whether restricted diffusion (‘brain’) scores derived from significant latent variables at baseline were associated with ED symptom severity scores at one-year follow-up, controlling for baseline ED severity scores.

### Supplementary analyses with PLS

We ran three supplementary analyses with PLS to see whether results of brain-behavior pattern mapping were specific to RND measures in ED patients. This included analyses with: i) fractional anisotropy MRI-derived features; ii) cortical thickness and subcortical volume features; and iii) brain (RND)-behavior patterns in controls. In all cases, behavioral measures remained the same, with the exception of the analysis in healthy controls, where we did not include ED symptom data.

## RESULTS

### Sample

Ninety-one adolescents meeting criteria for a DSM-V eating disorder (mean age = 16.1 years old, range = 13.1-18.2) were included. We also carried out a supplementary analysis with 48 healthy control (HC) volunteers (mean age = 16.2 years old, range =13.5–17.9). See **Table 1** for sample characteristics and descriptive statistics of included behavioral measures.

### Microstructural-behavioral patterns in adolescent eating disorders

Multivariate analysis with PLS identified two statistically significant latent variables (LVs) mapping patterns of behavior (including temperament, cognition, interoceptive awareness and emotion regulation) to age-corrected restricted diffusion across white matter tracts and subcortical structures (LV-1: permuted *p*<5e-8; LV-2: permuted *p*=0.035; **Figures 2A, 3A**; Supplementary Figure 1A). These two patterns explained 46.9% and 13.5% of the covariance, respectively, between restricted diffusion and behavior. We emphasize results from LV-1 in the main text, given the high degree of covariance LV-1 captures between brain and behavior, as well as its more reliable signature compared to LV-2, evidenced by out-of-sample cross validation analyses (see “*LV-1 brain-behavior correlations”* section below).

**Figure 2.**
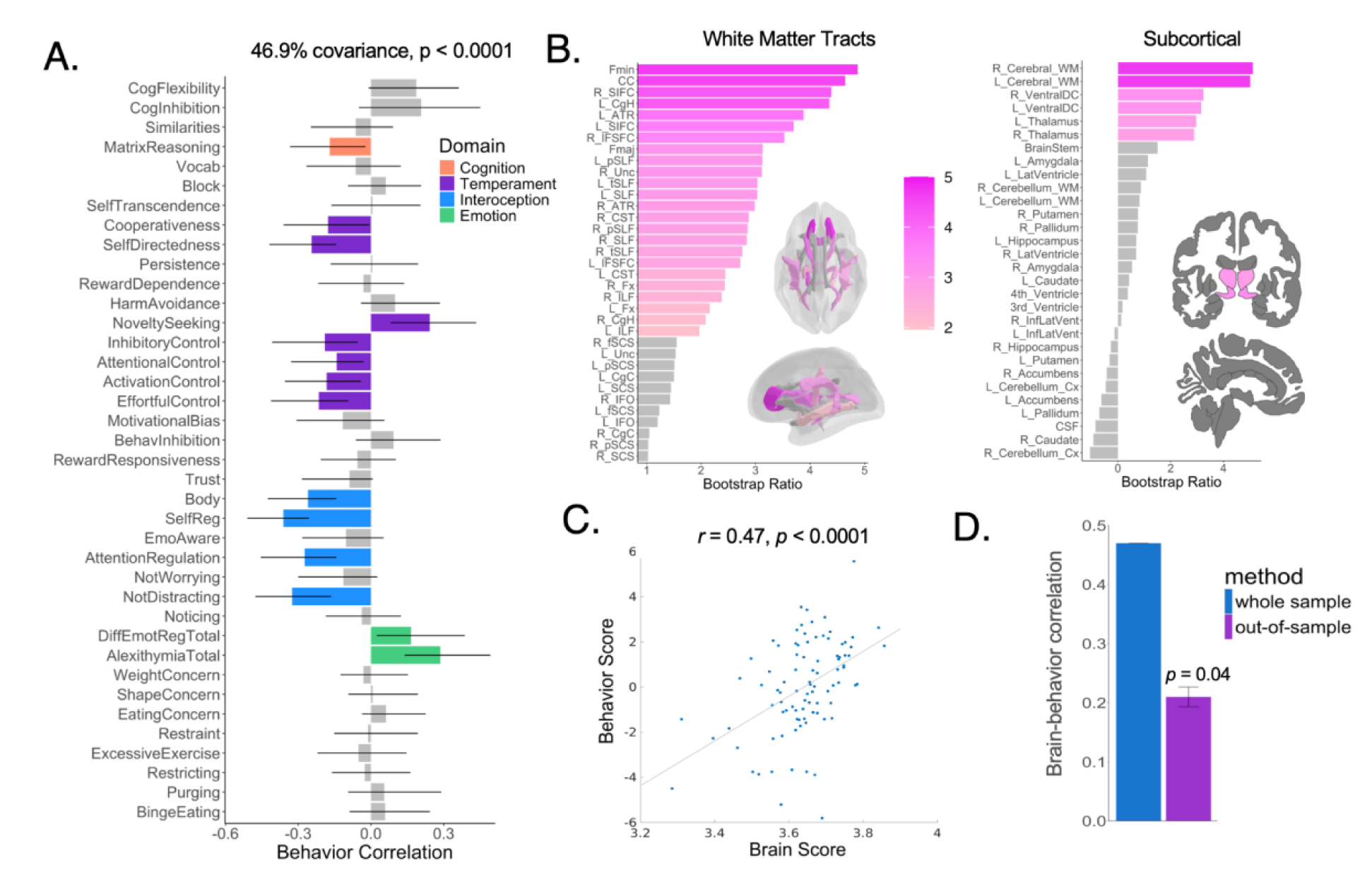
LV-1 results for brain measures derived from RND. <u>Panel A</u>: Behavioral loadings, shown with correlation coefficients, of each included behavioral measure on LV-1. Reliable loadings are color-coded by behavioral domain, where error bars indicate bootstrap-estimated standard errors. Loadings with error bars crossing zero were interpreted as non-significant loadings and are in grey. <u>Panel B</u>: The contribution of RND within individual brain ROIs to LV-1, plotted as bootstrap ratios (ratios between ROI weights and bootstrap-estimated standard errors), which can be interpreted as z-scores. The gradient depicts bootstrap ratios for regions with a bootstrap ratio > |1.96| (corresponding to 95% confidence interval), whereas ROIs falling under this threshold are grey. Note, adjacent brain maps visualized with *ggseg* omit some ROIs and are used for visualization purposes only. See Supplementary Tables 2 and 3 for ROI abbreviations. <u>Panel C</u>: The projection of individual patient data onto each of the weighted patterns in Panels A and B shows that brain and behavior scores are positively correlated. This suggests that patients who display the behavioral pattern in Panel A also tend to show increased RND in the significant brain regions in Panel B. <u>Panel D</u>: Correlations between brain-behavior scores in the full sample (same as Panel C) and in held-out data using the cross-validation scheme described in methods.

### LV-1 behavioral pattern

Figure 2A shows the loadings of each included behavioral measure to LV-1 (i.e., correlation coefficients between each behavioral variable and LV-1), indicating the contribution of a given behavioral measure. The strongest loadings emerged with interoceptive awareness, particularly with self-regulation (*r*=-0.36), where four of the seven included interoceptive measures reliably contributed to the LV-1 patterns based on bootstrap resampling. Seven of the 14 included temperament measures reliably contributed to LV-1 too, with the strongest associations emerging with lower self-directedness (*r*=-0.25) and higher novelty seeking (*r*=0.24). Other notable loadings included lower performance on matrix reasoning (*r=*-0.17) and more difficulties with emotion recognition (*r=*0.29) and regulation (*r*=0.17). ED symptom severity did not reliably contribute to LV-1.

### LV-1 microstructural pattern

Figure 2B shows the brain regions, across white matter tracts and subcortical structures, that contributed reliably to LV-1 as indexed by bootstrap ratios (95% confidence intervals). The majority of white matter tracts highly contributed to LV-1, suggesting a global contribution of white matter microstructure to LV-1, which was further confirmed by a high loading of cerebral white matter microstructure, defined by the *aseg* subcortical atlas. The strongest white matter tract loadings were found for the corpus callosum (including forceps major and minor) and bilaterally across frontal-striatal tracts, limbic tracts (parahippocampal portion of cingulum, fornix, uncinate fasciculus), and anterior thalamic radiations. For subcortical structures, microstructure of the ventral diencephalon and thalamus bilaterally contributed to LV-1.

### LV-1 Brain-behavior correlations

Figure 2C shows the correlation between individual behavioral and microstructural brain scores (*r*=0.47, p<0.0001). The mean out-of-sample correlation was assessed through cross validation with 100 randomized splits of the data into 75% train and 25% test (*r*=0.21, *p*=0.04) (Figure 2D).

### Summary of LV-2 brain-behavior results

Results for LV-2 with restricted diffusion are depicted in Figure 3. Behavioral loadings (Figure 3A) yielded a pattern of lower cognitive flexibility and inhibition (*r*∼-0.20), lower effortful control (*r*=0.15), and higher scores on self-transcendence, shape concern, and purging (*r*∼0.10). Interoceptive awareness and emotion-related measures did not contribute reliably to LV-2. Restricted diffusion measures revealed a contribution of various subcortical structures bilaterally, including basal ganglia, hippocampus, and lateral ventricles to LV-2 (Figure 3B). No white matter tracts contributed to LV-2. Although this LV was statistically significant after permutation testing (permuted *p*=0.035) and yielded significant brain-behavior score correlations (*r*=0.38*, p*<0.0001) (Figure 3C), we interpret this LV with caution given its low out-of-sample correlation performance (*r*=-0.035, *p*=0.52) (Figure 3D).

**Figure 3.**
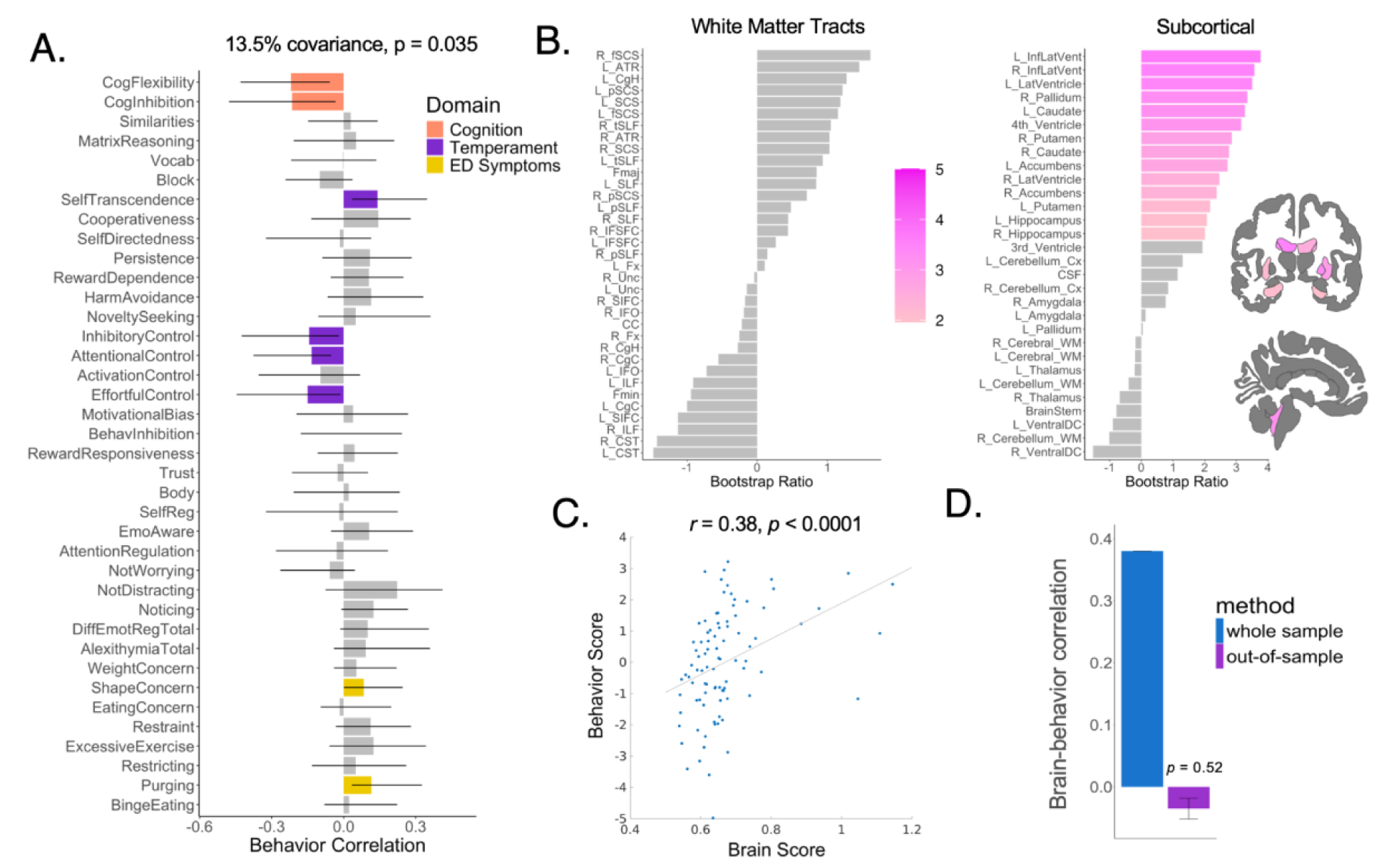
LV-2 results for brain measures derived from RND. <u>Panel A</u>: Behavioral loadings, shown with correlation coefficients, of each included behavioral measure on LV-1. Reliable loadings are color-coded by behavioral domain, where error bars indicate bootstrap-estimated standard errors. Loadings with error bars crossing zero were interpreted as non-significant loadings and are in grey. <u>Panel B</u>: The contribution of RND within individual brain ROIs to LV-1, plotted as bootstrap ratios (ratios between ROI weights and bootstrap-estimated standard errors), which can be interpreted as z-scores. The gradient depicts bootstrap ratios for regions with a bootstrap ratio > |1.96| (corresponding to 95% confidence interval), whereas ROIs falling under this threshold are grey. Note, adjacent brain maps visualized with *ggseg* omit some ROIs and are used for visualization purposes only. See Supplementary Tables 2 and 3 for ROI abbreviations. <u>Panel C</u>: The projection of individual patient data onto each of the weighted patterns in Panels A and B shows that brain and behavior scores are positively correlated. This suggests that patients who display the behavioral pattern in Panel A also tend to show increased RND in the significant brain regions in Panel B. <u>Panel D</u>: Correlations between brain-behavior scores in the full sample (same as Panel C) and in held-out data using the cross-validation scheme described in methods. Note, this LV needs to be interpreted with caution, given that it does not replicate in held-out data.

### LV-1 captures meaningful differences in ED subgroups

Next, we investigated whether the brain-behavior patterns identified by our significant LVs were driven more strongly by patients presenting with a binge-purge (*n*=30) vs. restricting (*n*=61) diagnostic subtype. For LV-1, relationships between individually-derived restricted diffusion and behavior scores were statistically significant across both groups (EDr: *r*=0.43, *p*=5.69e-4; EDbp: *r*=0.41, *p*=0.022) (Figure 4A). Similarly, brain-behavior correlations remained significant for both groups for LV-2 (EDr: *r*=0.40, *p*=1.29e-3; EDbp: *r*=0.50, *p*=5.10e-3). However, patients with the binge-purge subtype had higher scores across both LV-derived behavior (*t*(89)=-2.32, *p*=0.022) (Figure 4B) and restricted diffusion scores (*t*(89)=-3.66, *p*=4.26e-4) (Figure 4C). These group differences were not found for LV-2 (brain: *t*(89)=0.49, *p*=0.63; behavior: *t*(89)=0.81, *p*=0.42).

**Figure 4.**
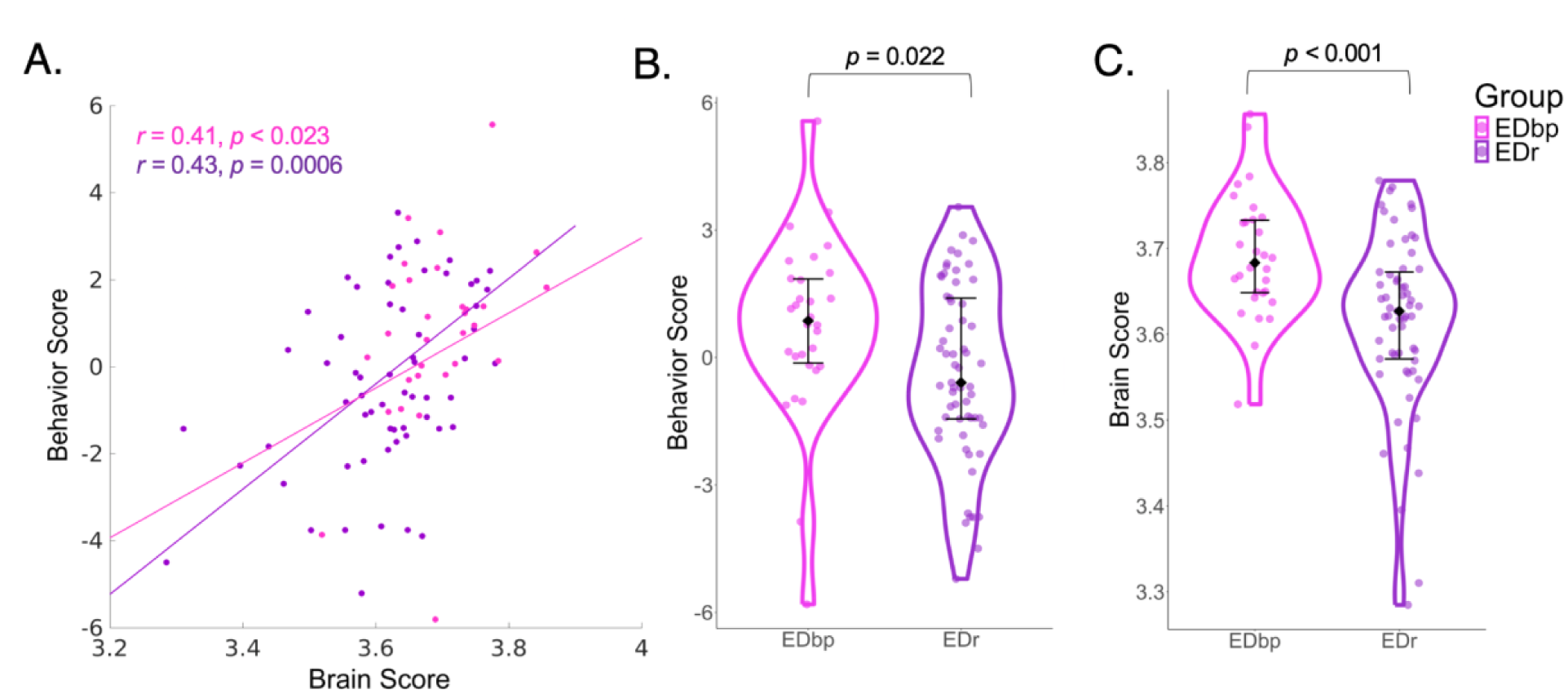
Stratifying LV-1 derived brain (RND) and behavior scores by patients with binge purge (EDbp; *n*=30) vs. restrictive (EDr; *n=*61) diagnostic subtype. <u>Panel A</u>: Brain-behavior correlations stratified by the two patient subgroups. Patient subgroup differences in behavior (<u>Panel B</u>) and brain scores (<u>Panel C</u>).

### Association of LV brain scores with ED symptom severity scores at one-year follow-up

We also investigated the potential prospective clinical utility of the uncovered restricted diffusion brain patterns, by associating LV-derived individual brain scores and ED symptom severity (assessed by global EDE-Q scores) one year later (*n*=72), controlling for baseline EDE-Q global scores. LV-2 derived brain scores, but not LV-1, were nominally associated with one-year ED symptom severity (*r*=0.25, *p=*0.03) (Figure 5).

**Figure 5.**
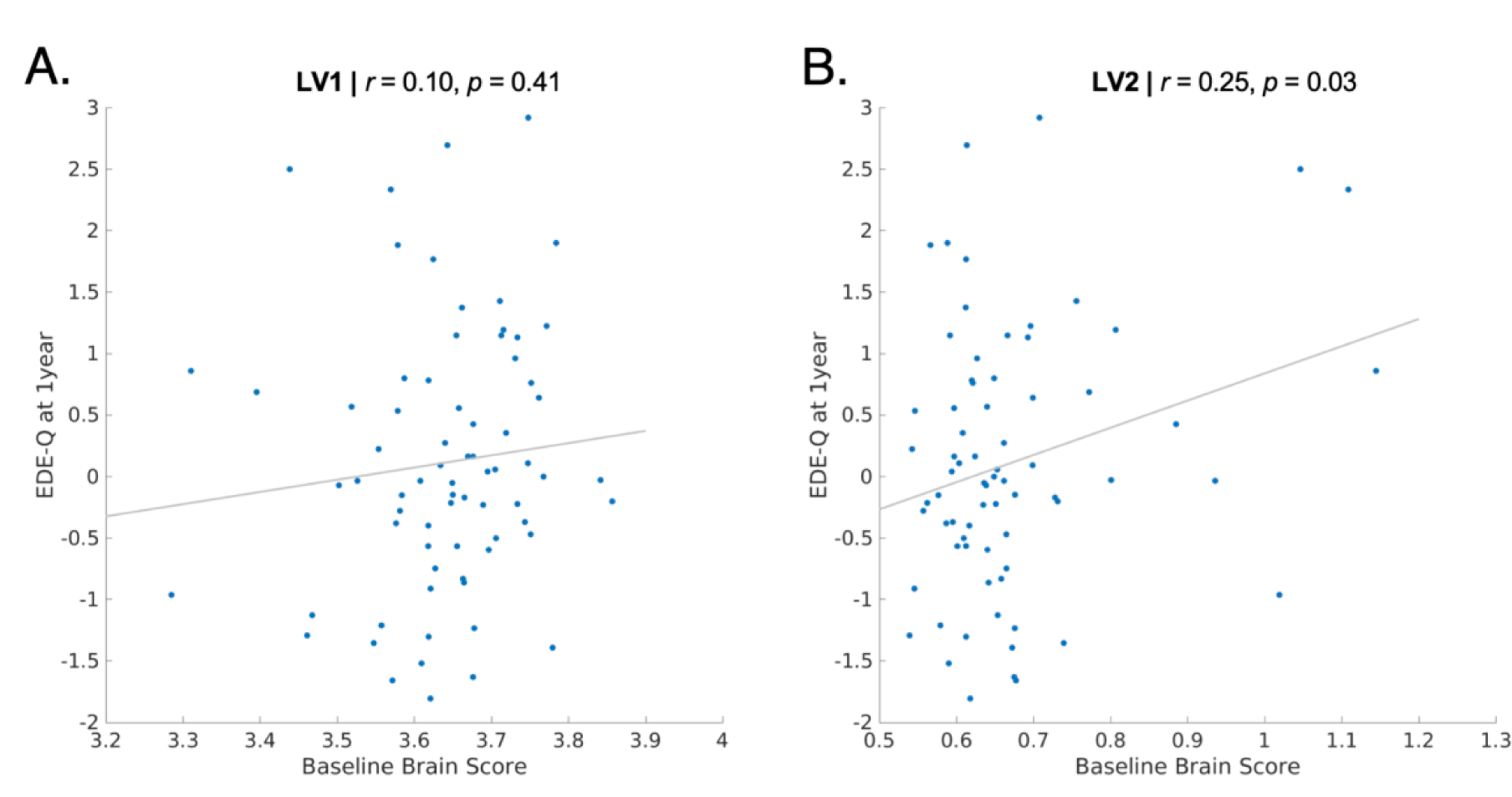
Brain scores (<u>Panel A</u>: LV-1; <u>Panel B</u>: LV-2), derived from PLS analysis with RND brain measures, and associations with EDE-Q scores one year later, controlling for baseline EDE-Q scores, for 72 patients with complete baseline and follow-up data.

### Specificity of LV-1 results to adolescent patients and restricted diffusion

Effect sizes for the first 15 LVs for all supplementary analyses can be found in Supplementary Figure 1. We observed a similar pattern of results when we used FA instead of RND (LV-1 in Supplementary Figure 2; LV-2 in Supplementary Figure 3). However, the two significant LVs with FA showed weaker associations compared to RND, most prominently for subcortical structures and ED symptoms for LV-2. Further, both LVs did not survive out-of-sample correlation tests with fractional anisotropy-derived brain measures (Supplementary Figures 2D, 3D). Similar to RND, FA-derived brain scores significantly differed between EDbp and EDr subgroups for LV-1 (*t*(89)=-3.22, *p*=1.80e-3), but not LV-2 (*t*(89)=-1.09, *p*=0.28). Unlike RND, LV-2 with FA measures was not significantly associated with one-year clinical symptoms (*r*=0.19, *p*=0.12) (Supplementary Figure 4). Although we found two significant LVs when mapping the same behavioral measures to cortical thickness and subcortical volumes (Supplementary Figure 1C), brain-behavior correlations in patients were not significant (Supplementary Figure 5), and did not survive out-of-sample cross-validation testing (LV1: out-of-sample *r*=-0.020, permuted *p*=0.53; LV2: out-of-sample *r*=-0.0051, permuted *p*=0.53). Finally, we did not find significant LVs when assessing restricted diffusion-behavior patterns in healthy controls (Supplementary Figure 1D).

## DISCUSSION

The current report harnesses a detailed collection of clinical, cognitive, and behavioral data in adolescent EDs with advanced diffusion imaging techniques, and reveals multivariate patterns across both brain and behavior that may capture clinically meaningful constructs. Our strongest multivariate associations were found with brain measures derived from restricted diffusion, where the first two latent variables explained 60.4% of the variance between brain and behavior. Notably, the first latent variable (46.9%) showed a significant out-of-sample brain-behavior correlation. Altogether, this first latent variable mapped a statistically robust behavioral profile of lower scores on abstract reasoning, effortful control, and interoceptive awareness, alongside higher emotional dysregulation and novelty seeking, to increased restricted diffusion across white matter tracts, particularly those joining frontal, limbic, and thalamic regions. This work emphasizes the power of using a multivariate and data-driven approach to showcase how distributed patterns of brain measures are collectively associated with different behavioral profiles in adolescent ED patients. Detection of such heterogeneity between neuroanatomical and behavioral patterns may otherwise have been missed with univariate statistics and a circumscribed focus on only a select few features within a ‘case-control’ framework; a framework that has often been adopted in ED research and other domains of psychiatric neuroimaging.

Brain measures derived from diffusion weighted imaging, particularly the RSI model, showed the strongest associations with behavior compared to structural morphometry measures. Given the similarity in derived latent variables with RND and FA, this work helps extend the interpretation of altered orientation of diffusion (as measured by FA) to intracellular tissue compartments (RND), likely reflecting differences in diffusion along neurites. LV-1 results with RND were replicated in out-of-sample correlation testing, which was not the case for results derived from FA. Finally, stronger patterns of associations arose with subcortical structures with RND compared to FA, particularly for LV-2. The observed pattern of increases in restricted diffusion, which was robustly correlated with an array of ED-related traits, may be driven by a number of biological processes. For instance, increases in RND can be observed with neuronal or glial cell death, increased lipid catabolism, shrinkage of glial cells, movement of intracellular fluid into extracellular spaces due to changes in oncotic pressure/dehydration, and/or alterations in endocrine signals (gonadal, thyroid, cortisol) [26, 39].

Importantly, the neurodevelopmental context of our sample needs to be considered in interpreting these findings. Microstructural markers are developing dynamically throughout childhood and adolescence, both within white matter and subcortical structures [31, 39], and can be shaped in response to environmental and activity-dependent experiences. Thus, diffusion imaging-based markers may be well-situated to study the neurobiology underlying ED-related behaviors in adolescence. In the current study, we found increased RND and FA in more anterior/frontal regions associated with more ED-related behaviors. Of developmental relevance, increases in these microstructural markers are often associated positively with age in adolescence, with some evidence for a posterior to anterior gradient across development in childhood and adolescence [88]. It is possible that our findings, particularly for frontal and anterior subcortical microstructure, may reflect an accelerated maturation in microstructure in ED patients, and in turn, contributing to differences in emotion regulation, interoceptive awareness, abstract reasoning and temperamental traits compared to healthy controls. We did not find any robust brain-behavior links with structural MRI markers (cortical thickness, subcortical volumes); it is possible that these imaging measures are better at distinguishing broad case-control differences [27, 53] rather than the contribution of brain architecture to heterogeneous behaviors in adolescence. We controlled for age in the current analyses, thus further work mapping deviations in neurodevelopmental trajectories in EDs from typically developing adolescents are warranted.

The highest behavioral loadings for LV-1 included measures of interoceptive awareness and temperament, particularly lower self-regulation, which reflects a poorer ability to regulate psychological distress through focusing attention on body sensations [89]. Lower effortful control, emotion recognition and regulation, poor abstract reasoning, and high novelty seeking also contributed to this behavioral profile. Altogether, this pattern parallels that of an ‘undercontrolled’ or dysregulated personality profile that has been described in temperament-based clustering of ED patients [15, 90, 91] and is more often observed in ED patients with binge and/or purge symptoms [16]. Our results show that patients with a binge-purge diagnostic subtype have both higher brain and behavior LV-scores compared to the ED-restricting subtype. It should be noted, however, that this latent variable still captured significant brain-behavior associations in each subtype individually, suggesting that this brain-behavior pattern is capturing a transdiagnostic phenotype. Further, the high covariance explained between microstructure and ‘undercontrolled’ behavior in LV-1 may be reflecting the strong biological basis of uncovered traits (e.g., temperament, interoceptive awareness). This is further supported by previous work in our group, where we found evidence for shared genetic architecture between restricted diffusion measures across the brain and risk-taking behaviors [92].

We also found a significant positive association between higher restricted diffusion across subcortical structures, as indexed by LV-2 brain scores at baseline, and higher ED symptoms one year later. However, this association did not survive correction for multiple comparisons, and these brain scores were derived from a latent variable that did not replicate in our cross-validation scheme. It should also be noted that there are many factors that are contributing to a patient’s ED symptoms after one year that were not accounted for in this study. It remains important for future studies to capture more dynamic changes in both brain and behavior, particularly in efforts to predict symptoms that rapidly fluctuate over the course of the illness, such as disordered eating behaviors. Temperament-based measures also have the capacity to change over the course of treatment; for instance, our group has previously found that changes in motivational bias rather than just baseline reward/punishment sensitivity was predictive of clinical symptoms at discharge [93]. Restricted diffusion is also notably changing during adolescence [39], and prediction of clinical symptoms may benefit from inclusion of neurodevelopmental trajectories, as opposed to a single imaging time point.

Other factors should be considered when interpreting this work. First, only female participants were included. EDs among males are relatively rare [94] and have a different clinical presentation [95, 96]. Our work contributes to a growing emphasis on the importance of prioritizing efforts to understand women’s health and sex differences in mental health [97, 98]. Opportunities in ongoing and emerging large-scale data collection efforts will also help address brain-behavior associations in adolescent males compared to females with EDs, such as the Adolescent Brain Cognitive Development^SM^ (ABCD^®^) Study. We also included a relatively small number of healthy controls, which could explain why we did not find any significant latent variables when mapping brain and behavior in this sample. Although our included sample of ED patients was better powered than many previously published neuroimaging studies, a larger sample would be required to look at more fine-grained neuroimaging patterns across the brain, beyond the pre-defined regions of interest included in the current investigation. However, we included all regions of interest available through the included atlases to maintain a data-driven approach. Finally, our sample included patients with various ED diagnoses, including patients with avoidant/restrictive food intake disorder (ARFID), purging disorder and other specified feeding disorder (OSFED). We believe that our chosen multivariate approach helps address this heterogeneity, given that our goal was not to distinguish patients based on diagnostic category, but rather to characterize behavioral and brain-based patterns transdiagnostically across EDs in adolescence.

Altogether a multivariate approach captured a high degree of covariance (46.9%) between microstructure connecting frontal, limbic and thalamic regions, and temperament profiles in adolescent patients with an eating disorder. Our results emphasize the value of applying multivariate methods to more accurately and reproducibly model the array of brain-behavior relationships inherent to the heterogeneous clinical presentation of eating disorders. These behavioral and brain-based patterns may also be important in differentiating between clinical subgroups of patients and in future prediction-based frameworks of clinical outcomes in eating disorders.

## ACKNOWLEDGMENTS

The authors would like to thank all participants who were involved in the study. The authors would also like to thank Miki Carrico for initial quality control of the imaging data. Dr. Anders M. Dale is Founding Director, holds equity in CorTechs Labs, Inc. (DBA Cortechs.ai), and serves on its Board of Directors and the Scientific Advisory Board. He is an unpaid consultant for Oslo University Hospital. The terms of these arrangements have been reviewed and approved by the University of California, San Diego in accordance with its conflict-of-interest policies. All other authors have no conflicts of interest.

## AUTHOR CONTRIBUTIONS

CM, ABG, and CEW contributed to the design of the study. MM, CFN, ABG and CEW contributed to collection and curation of the data. CM, DJH, DP, and AMD contributed to image processing through restriction spectrum imaging. CM and GS wrote the code to analyze and visualize the data. CEW and AMD provided mentorship for the project. CM, CEW, and ABG provided funding for the study. All authors contributed to interpretation of the results, and reviewed and provided feedback on the manuscript.

## DATA AVAILABILITY STATEMENT

The data included in this manuscript is available on the NIH Data Archive (NDA), under the NDA Study “Neurocircuitry of Temperament and Motivated Behavior in Adolescent Eating Disorders” (#2736).

