## Supplementary for "Multivariate patterns linking brain microstructure to temperament and behavior in adolescent eating disorders"

[1]Center for Multimodal Imaging and Genetics, University of California San Diego, La Jolla, California, USA
[2]Department of Radiology, University of California San Diego, La Jolla, California, USA
[3]Department of Psychiatry, Perelman School of Medicine, University of Pennsylvania, Philadelphia, Pennsylvania
[4]Department of Psychiatry, University of California San Diego, La Jolla, California, USA
[5]Department of Neurosciences, University of California San Diego, La Jolla, California, USA
*Authors share senior authorship.

**Sample recruitment & inclusion criteria**

Female adolescents with an ED, meeting criteria for a DSM-V restricting and binge-purge type eating disorder [1, 2], were recruited from the University of California, San Diego Eating Disorders Treatment and Research Program. ED diagnosis was determined by meeting *DSM-V* criteria through the Kiddie Schedule for Affective Disorders and Schizophrenia (KSADS-5) [3, 4], a semi-structured interview performed by a trained research assistant, under the supervision of a doctoral level psychologist. HCs were recruited from the San Diego community. See Table 1 of the main manuscript for diagnostic breakdown of the patient sample.

Exclusion criteria across patients with an eating disorder and healthy controls (HC) included: history of alcohol or drug abuse or dependence 3 months prior to study, medical or neurological concerns including a history of head injury with loss of consciousness, and intellectual or developmental disability. Individuals were not excluded if they were taking psychoactive medications. HCs were additionally excluded if they had any eating disorder symptoms, as determined by the Eating Disorders Examination (EDE) [5, 6], and/or a history of an eating disorder or any other Axis I disorder, as determined by the KSADS-5 [3, 4]. All study procedures were approved by the University of California, San Diego's Institutional Review Board (170664) and informed consent/assent was obtained prior to initiation of study procedures.

**MRI Acquisition.**

The imaging visit was scheduled during the early follicular phase (days 1 to 10) of the subject's menstrual cycle, if she had begun (or resumed) menses. The imaging visit was conducted in the morning after an overnight fast, given the broader study protocol which included incentive processing of different taste stimuli. Self-report assessments relevant to this study (see Supplementary Table 1) were completed after the scan.

MRI Images were collected on a 3.0 T GE MR750 scanner equipped with quantum gradients providing echo planar capability, using a Nova Medical 32 channel head coil (maximum gradient strength: 50 mT/m, slew rate: 200 T/m/s). The following protocol was administered to acquire structural T1-weighted images and multi-shell diffusion: a three-plane localizer scan; a whole brain, sagittally acquired (0.8 mm slice thickness, FOV=256 mm) T1-weighted (MPRAGE PROMO, TE=3.656 ms, flip angle=$8^o$, matrix=320, 2x in-plane acceleration) and separate T2-weighted (3D CUBE, 0.8 mm slice thickness, FOV= 256 mm, TE=60 ms, variable flip angle, matrix=320, 2x in-plane acceleration) sequence for alignment and morphometry; and two DTI scans (FOV=240 mm, slice thickness = 1.7 mm, matrix = 140 x 140, b=1500/3000 s/$mm^2$; 102 diffusion directions) which were further used for restriction spectrum imaging (RSI) modeling, detailed in the section below.

*Image Quality Control*. During the scan, visual inspection of the images were combined with real time image reconstruction and online calculation of QC metrics. When compromised image quality was detected, the scan was stopped, and additional instructions were communicated with the participant to ensure compliance. After acquisition, images further underwent quality control procedures, adapted by the Human Connectome Project protocol. This included: 1) visual rating of the T1- and T2-weighted images; 2) visual assessment of DTI distortion; and 3) detection of motion artifacts. One hundred and five patients with an ED and 50 HC completed an MRI scan at baseline. Of this sample, 6 ED patients were not further processed due to failing the above quality control measures.

**MRI Processing Pipeline.**

Data were processed using the Adolescent Brain Cognitive Development$^{SM}$ (ABCD$^{®}$) Study pipeline for images acquired on a GE MR750 scanner, as described in [7]. All imaging measures relied on collection of T1-weighted (T1w) structural MRI images (1mm isotropic), which were acquired with a 3D T1w inversion prepared RF-spoiled gradient echo scan. After processing, an additional ED patient and 2 healthy controls failed the pipeline, leaving a final sample size of 98 patients and 48 healthy controls with high quality imaging data.

*Modeling of diffusion-weighted data with Restriction Spectrum Imaging (RSI).* We used RSI to model restricted directional diffusion (RND) across the entire brain. Details of this approach can be found in previous publications [8–10]. Briefly, RSI takes advantage of a multi-shell diffusion acquisition to estimate the contribution of diffusion signal from separable pools of water within a tissue, which includes free water (e.g., CSF), hindered diffusion (e.g., mostly extracellular space), and restricted diffusion (e.g., mostly intracellular space). Of interest to this study, restricted diffusion describes water within intracellular spaces confined by cell membranes with a non-Gaussian pattern of displacement. Spherical deconvolution (SD) is used to reconstruct the fiber orientation distribution (FOD) in each voxel from the restricted compartment, where the restricted tissue compartment is modeled as a fourth order spherical harmonic (SH) function. The restricted directional measure, RND, is the norm of the SH coefficients for the second and fourth order SH coefficients (divided by the norm of the coefficients across restricted, hindered, and free water compartments). In other words, RND models diffusion emanating from multiple directions within a voxel.

As a comparison to RND within superficial white matter, we also included fractional anisotropy (FA), derived from the diffusion tensor model [11, 12]. Diffusion tensor parameters were calculated using a standard, linear estimation approach with log-transformed diffusion-weighted (DW) signals [12]. Tensor matrices were diagonalized using singular value decomposition, obtaining three eigenvectors and three corresponding eigenvalues, from which FA could be calculated [11].

*Cortical thickness and volumes*. Cortical surfaces were constructed from T1-weighted structural images for each subject and segmented to calculate measures of cortical thickness along 5124 vertices using FreeSurfer v7.1.1 [13–17]. Cortical maps were smoothed using a Gaussian kernel of 20 mm full-width half maximum (FWHM) and mapped into standardized spherical atlas space. Processing with Freesurfer also yields automated tissue segmentation [18], which was used to determine volumes of subcortical structures (see section below).

**MRI-derived regions of interest.**

See Supplementary Tables 2-4 for a full list of ROIs used. For RND and FA analyses, 65 ROIs were used across 35 white matter tracts from the *AtlasTrack* probabilistic atlas [19] (Supplementary Table 2) and 30 subcortical structures from Freesurfer's *aseg* atlas [18] (Supplementary Table 3). For morphometric analysis, cortical thickness across 68 regions from the *Desikan-Killiany* atlas [20] (Supplementary Table 4) and volumes for the above-mentioned 30 subcortical structures were included. Note, we retained ventricles and cerebrospinal fluid ROIs for RSI measures because it is possible to have restricted diffusion in ventricles if inflammatory markers are present. For instance, cerebral abscesses have been noted in patients with AN [21], which can cause diffuse patterns of abnormal restricted diffusion [22].

**Behavior**

Please see Supplementary Table 1 below for the behavioral instruments and variables included in analysis. Seven ED patients were dropped due to missing behavioral data in one or more of the instruments, resulting in our final sample reported in the main manuscript of 91 patients and 48 HC.

**Partial Least Squares model**

Partial Least Squares (PLS) is a form of reduced rank regression that identifies linear combinations of two sets of variables that maximally covary with each other [23, 24]. In the present study, one set represents neuroimaging-derived data for ED patients (denoted as $\mathbf{X}_{nxp}$), while the other set corresponds to the behavioral measures (denoted as $\mathbf{Y}_{nxq}$). The *n* rows of both matrices **X** and **Y** represent the number of individuals with an ED (i.e., *n*=91). The *p* columns of matrix **X** correspond to the number of brain measures. For diffusion-based ROIs, we included 65 regions (35 white matter tracts, 30 subcortical structures); for the supplementary analyses with cortical thickness and volumes, 98 regions were included (68 cortical, 30 subcortical). The values in matrix **X** were then corrected for age using a linear regression model. The *q* columns of matrix **Y** correspond to the behavioral measures from the same ED patients, including 38 measures that capture clinical symptoms, cognition, temperament, and interoceptive awareness (See Supplementary Table 1 below). Finally, both **X** and **Y** matrices were standardized column-wise (i.e., z-scored) and a correlation matrix (**X'Y**) was computed from the standardized matrices. Singular value decomposition (SVD) was then applied to the correlation matrix **R=X'Y** as follows:

$$\mathbf{R}=\mathbf{X}'\mathbf{Y} = \mathbf{U}\mathbf{S}\mathbf{V}'$$

The decomposition results in two orthonormal matrices of left and right singular vectors (**U** and **V**, respectively), and a diagonal matrix of singular values (**S**). The main results of PLS analysis are latent variables, which are weighted linear combinations of the original variables from the two initial variable sets (i.e., **X** and **Y**). PLS latent variables are mutually orthogonal and express the shared information between the two sets with maximum covariance. Latent variable *i* ($LV_i$) is composed of the *i*th column vector of **U**, *i*th column vector of **V**, and the *i*th singular value from **S**. The elements of the column vectors of **U** and **V** are the weights of the original neuroimaging-derived values and behavioral measures, respectively, that contribute to the latent variable. The covariance between neuroimaging-derived and behavioral patterns is reflected in the corresponding singular values from the diagonal elements of matrix **S** and is estimated for the latent variable *i* as follows:

$$\eta_i = \frac{s_i^2}{\sum_{j=1}^{J} s_j^2}$$

where $\eta_i$ is the effect size for $LV_i$, $s_i$ is the corresponding singular value from the diagonal matrix **S**, and *j* is the total number of singular values. Furthermore, the PLS-derived neuroimaging and behavioral patterns (i.e., left and right singular vectors, **U** and **V**) can be used to estimate patient-specific scores that reflect how much each patient expresses the PLS-derived patterns. The patient-specific brain and behavioral scores are calculated by projecting the neuroimaging and behavioral patterns (i.e., **U** and **V**) onto the original patient data:

Brain score = **XU**

Behavioral score = **YV**

We performed three additional analyses to assess the significance, reliability, and generalizability of the findings: (a) permutation testing to assess the statistical significance of the overall patterns; (b) bootstrap resampling to assess feature (imaging, behavioral) importance; (c) cross-validation analysis to assess the out-of-sample correlations between projected scores. Each step is discussed below.

*Permutation tests*. We assessed the statistical significance of each latent variable using permutation tests[63] by randomizing the correspondence between brain and behavioral measures Specifically, PLS analysis was repeated after randomly reordering the rows of variable set **X** during each permutation. The procedure was repeated 10,000 times resulting in a null distribution of singular values. To test the null hypothesis that there is no relationship between imaging and behavioral measures, a *p*-value was estimated for each latent variable as the proportion of the times that the permuted singular values were greater than or equal to the original singular value.

*Bootstrap resampling*. The reliability of singular vector weights (i.e., weights of neuroimaging-derived values and behavioral variables) were assessed using bootstrap resampling (10,000 repetitions)[64]. The rows of the two variable sets (i.e., **X** and **Y**) were randomly resampled with replacement and PLS analysis was repeated with the new correlation matrix for each bootstrapped sample. This generated a sampling distribution for each neuroimaging and behavioral weight. To assess the reliability of each variable, bootstrap ratios were calculated as the ratio of each variable's weight to its bootstrap-estimated standard error. Bootstrap ratios can be used to identify variables (brain regions of interest or behavioral measures) that make a large contribution to the overall pattern (i.e. have a large weight) and, at the same time, are stable across individuals (i.e. have a small standard error). If the bootstrap distribution is Gaussian, a bootstrap ratio can be interpreted as a z-score[64], such that 95% and 99% confidence intervals correspond to bootstrap ratios of $\pm$1.96 and $\pm$ 2.58, respectively.

*Cross-validation*. Cross-validation analysis was used to assess the out-of-sample correlation between brain and behavior scores[65,110]. We used 100 randomized train-test splits of the original data, where 75% of the data was treated as a training set and 25% of the data was treated as an out-of-sample test set. For each training set, PLS was used to estimate neuroimaging-derived and behavioral patterns (i.e., $\mathbf{U}_{train}$ and $\mathbf{V}_{train}$). The test data were then projected onto the neuroimaging-derived and behavioral patterns derived from the training set, to estimate patient-specific scores and their correlation for the test sample (i.e. $corr(\mathbf{X}_{test}\mathbf{U}_{train}, \mathbf{Y}_{test}\mathbf{V}_{train})$) . This procedure was repeated 100 times to generate a distribution of out-of-sample correlation coefficients. Finally, we used permutation tests (100 repetitions) to assess the significance of these out-of-sample correlation coefficients, where we randomly reordered rows of the original neuroimaging matrix and repeated the above procedure for each permutation. The procedure generated a null distribution of correlation coefficients between neuroimaging and behavioral scores in the test sample. A *p*-value was then calculated as the proportion of correlation coefficients that were greater than or equal to the mean original out-of-sample correlation coefficient.

| Domain | Instrument | Variables |
|---|---|---|
| Cognition | Wechsler Abbreviated Scale of Intelligence (WASI) [25] | Similarities raw score<br>Matrix Reasoning raw score<br>Vocabulary (Vocab) raw score<br>Block Design (Block) raw score |
| | Flanker task [26] | Flanker uncorrected score (Cognitive inhibition or "CogInhibition" in manuscript) |
| | NIH Toolbox, Dimensional Change Card Sort [27] | Dimensional Change Card Sort, uncorrected score (Cognitive flexibility or "CogFlexibility" in manuscript) |
| Temperament | Temperament and Character Inventory (TCI) [28] | Self Transcendence<br>Cooperativeness<br>Self Directedness<br>Persistence<br>Reward Dependence<br>Harm Avoidance<br>Novelty Seeking |
| | The Adult Temperament Questionnaire (ATQ) [29] | Inhibitory Control<br>Attentional Control<br>Activation Control<br>Effortful Control |
| | Behavioral Inhibition and Behavioral Activation Scales (BISBAS)[30] | Behavioral Inhibition<br>Reward Responsiveness<br>Motivational Bias |
| Interoceptive awareness | Multidimensional Assessment of Interoceptive Awareness (MAIA) [31] | Trust<br>Body<br>Self Regulation (Self Reg)<br>Emotional Awareness (Emo Aware)<br>Attention Regulation<br>Not Worrying<br>Not Distracting<br>Noticing |
| ED symptoms | Eating Disorders Examination (EDE) [5, 6] | Weight Concern<br>Shape Concern<br>Eating Concern<br>Restraint |
| | Eating Pathology Symptoms Inventory (EPSI) [32] | Excessive Exercise<br>Restricting<br>Purging<br>Binge Eating |

| | | |
|---|---|---|
| Emotion | Difficulties in Emotion Regulation Scale (DERS) [33] | Difficulties in Emotion Regulation (Diff Emot Reg) Total Score |
| | Toronto Alexithymia Scale (TAS) [34] | Alexithymia Total Score |

**Supplementary Table 1**. Behavioral and clinical measures. All variables reflect standard subscale scores obtained directly from the instrument, with the exception of Motivational Bias from the BISBAS. Motivational bias was calculated as the difference between reward responsiveness and inhibition, reflecting the dominance of reward vs punishment sensitivity [35]. Some subscales from the EPSI were omitted as they were already largely captured by the EDE (cognitive restraint, body dissatisfaction), and/or have not been shown to have a significant bearing on clinical course in adolescent girls (muscle building, obesity attitude).

| Abbrev | Fiber Name | Connected Brain Regions |
|---|---|---|
| Fx | fornix | hippocampus & mammillary nuclei of hypothalamus |
| CgC | cingulate cingulum | cingulate gyrus & entorhinal cortex (cingulate portion) |
| CgH | parahippocampal cingulum | cingulate gyrus & entorhinal cortex (parahippocampal portion) |
| CST | corticospinal tract (pyramidal tract) | motor cortex & spinal cord |
| ATR | anterior thalamic radiations | thalamus & frontal lobe |
| UNC | uncinate | inferior frontal lobe & anterior temporal lobe |
| ILF | inferior longitudinal fasciculus | occipital lobe & temporal lobe |
| IFO | inferior frontal occipital fasciculus | occipital lobe & frontal lobe |
| Fmaj | forceps major | left occipital cortex & right occipital cortex |
| Fmin | forceps minor | left prefrontal cortex & right prefrontal cortex |
| CC | corpus callosum | left cortex & right cortex |
| SLF | superior longitudinal fasciculus | temporal and parietal lobes & frontal lobe |
| tSLF | temporal superior longitudinal fasciculus (arcuate fasciculus) | temporal lobe & frontal lobe |
| pSLF | parietal superior longitudinal fasciculus | parietal lobe & frontal lobe |
| SCS | superior corticostriate | superior cortex & striatum |
| fSCS | frontal superior corticostriate | superior frontal cortex & striatum |
| pSCS | parietal superior corticostriate | superior parietal cortex & striatum |
| SIFC | striatal inferior frontal cortex tract | inferior frontal cortex & striatum |
| IFSFC | inferior frontal to superior frontal cortical tract | inferior frontal cortex & superior frontal cortex |

**Supplementary Table 2**. White matter tract regions of interest, available through the *Atlas Track* parcellation [7, 19], used for analyses with restricted normalized diffusion (RND) and fractional anisotropy (FA). All ROIs have separate labels for left and right hemispheres, except for corpus callosum, forceps major and forceps minor.

| ROI name | Description |
|---|---|
| Cerebral_WM | Cerebral white matter |
| Lateral-Ventricle | Lateral ventricle |
| Inf-Lat-Vent | Inferior lateral ventricle |
| Cerebellum-WM | Cerebellum white matter |
| Cerebellum-Cx | Cerebellum grey matter |
| Thalamus | Thalamus-proper |
| Caudate | Caudate |
| Putamen | Putamen |
| Pallidum | Pallidum |
| Hippocampus | Hippocampus |
| Amygdala | Amygdala |
| Accumbens | Accumbens Area |
| VentralDC | Ventral diencephalon |
| 3rd_Ventricle | Third ventricle |
| 4th_Ventricle | Fourth ventricle |
| Brain_Stem | Brain stem |
| CSF | Cerebrospinal fluid |

**Supplementary Table 3**. Subcortical regions of interest, available through the *aseg* parcellation in Freesurfer (Fischl et al., 2002), and used in analyses of RND, FA, and volumes. All structures are labeled on both hemispheres, except for the third and fourth ventricles, brainstem, and cerebrospinal fluid.

| ROI name | Description |
|---|---|
| bankssts | Banks superior temporal sulcus |
| caudalanteriorcingulate | Caudal anterior-cingulate cortex |
| caudalmiddlefrontal | Caudal middle frontal gyrus |
| cuneus | Cuneus cortex |
| entorhinal | Entorhinal cortex |
| fusiform | Fusiform gyrus |
| inferiorparietal | Inferior parietal cortex |
| inferiortemporal | Inferior temporal gyrus |
| isthmuscingulate | Isthmus-cingulate cortex |
| lateraloccipital | Lateral occipital cortex |
| lateralorbitofrontal | Lateral orbital frontal cortex |
| lingual | Lingual gyrus |
| medialorbitofrontal | Medial orbital frontal cortex |
| middletemporal | Middle temporal gyrus |
| parahippocampal | Parahippocampal gyrus |
| paracentral | Paracentral lobule |
| parsopercularis | Pars opercularis |
| parsorbitalis | Pars orbitalis |
| parstriangularis | Pars triangularis |
| pericalcarine | Pericalcarine cortex |
| postcentral | Postcentral gyrus |
| posteriorcingulate | Posterior-cingulate cortex |
| precentral | Precentral gyrus |
| precuneus | Precuneus cortex |
| rostralanteriorcingulate | Rostral anterior cingulate cortex |
| rostralmiddlefrontal | Rostral middle frontal gyrus |
| superiorfrontal | Superior frontal gyrus |
| superiorparietal | Superior parietal cortex |
| superiortemporal | Superior temporal gyrus |
| supramarginal | Supramarginal gyrus |
| frontalpole | Frontal pole |
| temporalpole | Temporal pole |
| transversetemporal | Transverse temporal cortex |
| insula | Insular cortex |

**Supplementary Table 4**. Cortical regions of interest, available through the *Desikan-Killiany* parcellation [20] and used in supplementary analyses with cortical thickness. All structures are labeled on both left and right hemispheres.

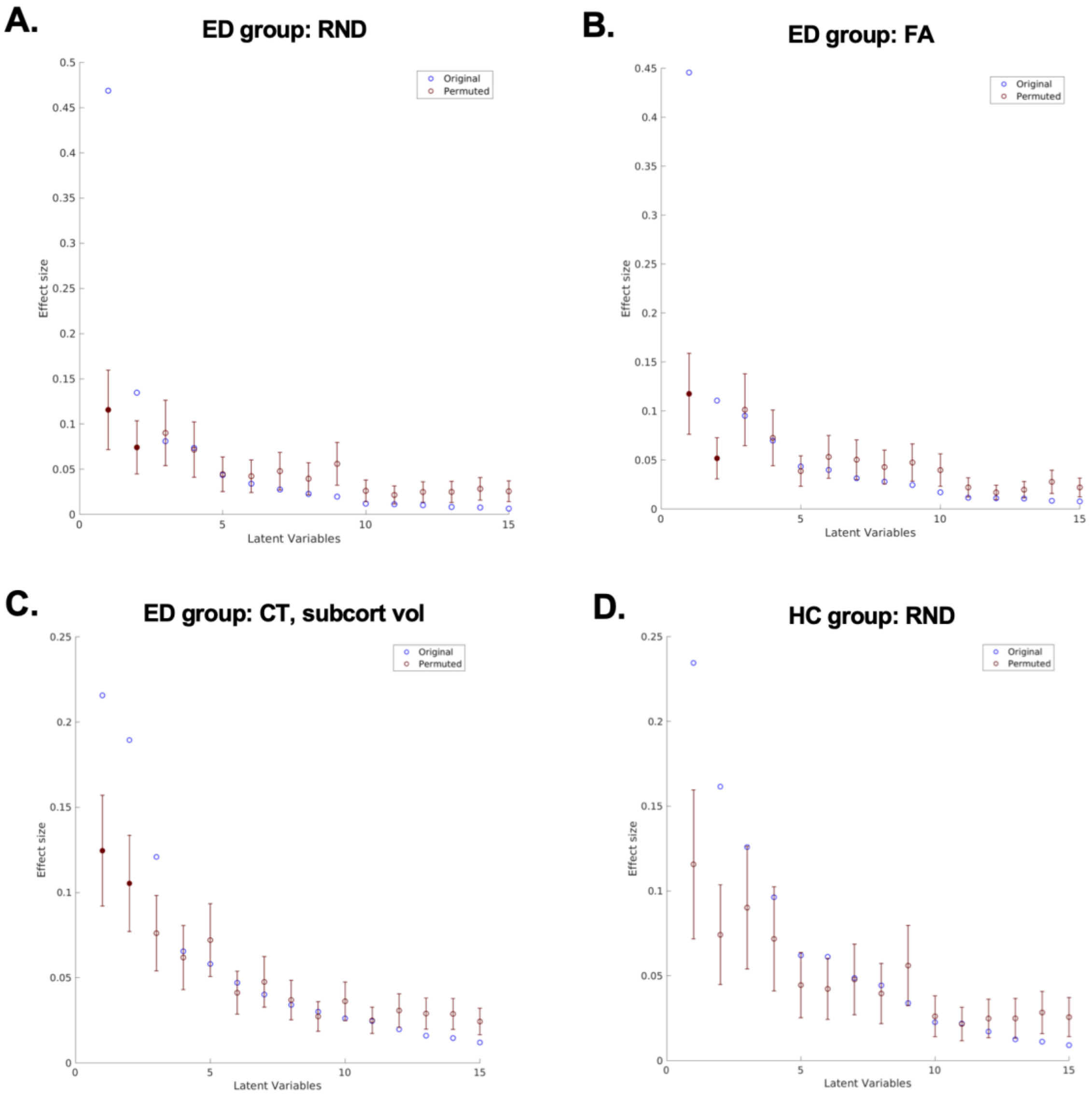


**Supplementary Figure 1**. Effect sizes (variance explained) across the top 15 LVs derived for each PLS analysis investigated in the current study. Filled red dots denote significant LVs, with a permuted *p*-value<0.05.

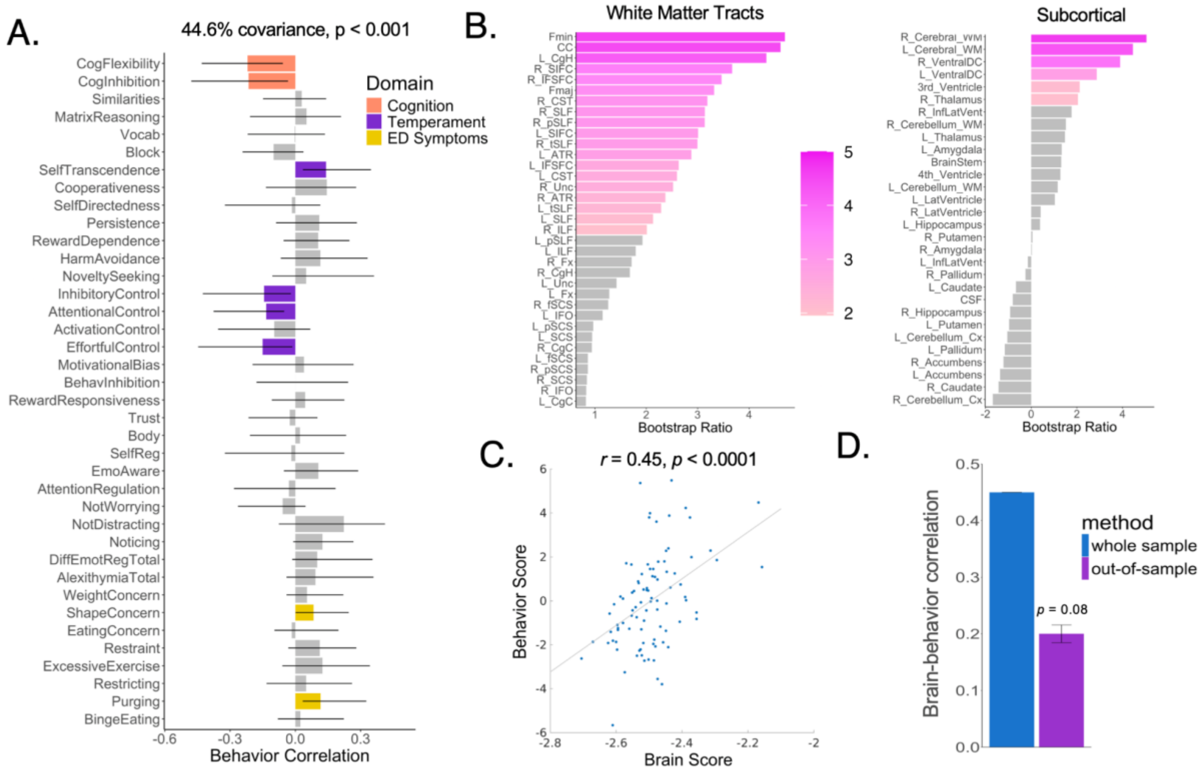


**Supplementary Figure 2.** LV-1 results for brain measures derived from FA. Panel A: Behavioral loadings, shown with correlation coefficients, of each included behavioral measure on LV-1. Reliable loadings are color-coded by behavioral domain, where error bars indicate bootstrap-estimated standard errors. Loadings with error bars crossing zero were interpreted as non-significant loadings and are in grey. Panel B: The contribution of RND within individual brain ROIs to LV-1, plotted as bootstrap ratios (ratios between ROI weights and bootstrap-estimated standard errors), which can be interpreted as z-scores. The gradient depicts bootstrap ratios for regions with a bootstrap ratio > |1.96| (corresponding to 95% confidence interval), whereas ROIs falling under this threshold are grey. See Supplementary Tables 2 and 3 for ROI abbreviations. Panel C: The projection of individual patient data onto each of the weighted patterns in Panels A and B shows that brain and behavior scores are positively correlated. This suggests that patients who display the behavioral pattern in Panel A also tend to show increased FA in the significant brain regions in Panel B. Panel D: Correlations between brain-behavior scores in the full sample (same as Panel C) and in held-out data using the cross-validation scheme described in methods.

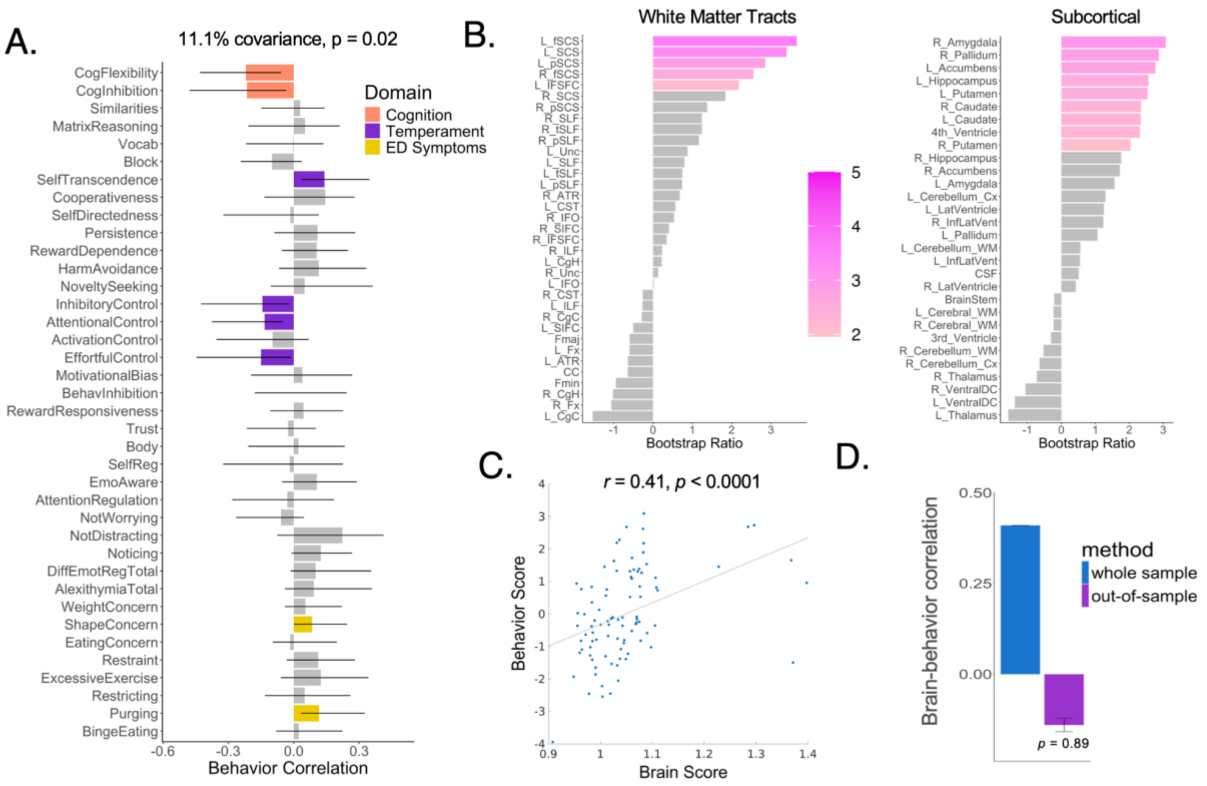


**Supplementary Figure 3.** LV-2 results for brain measures derived from FA. Panel A: Behavioral loadings, shown with correlation coefficients, of each included behavioral measure on LV-1. Reliable loadings are color-coded by behavioral domain, where error bars indicate bootstrap-estimated standard errors. Loadings with error bars crossing zero were interpreted as non-significant loadings and are in grey. Panel B: The contribution of RND within individual brain ROIs to LV-1, plotted as bootstrap ratios (ratios between ROI weights and bootstrap-estimated standard errors), which can be interpreted as z-scores. The gradient depicts bootstrap ratios for regions with a bootstrap ratio > |1.96| (corresponding to 95% confidence interval), whereas ROIs falling under this threshold are grey. See Supplementary Tables 2 and 3 for ROI abbreviations. Panel C: The projection of individual patient data onto each of the weighted patterns in Panels A and B shows that brain and behavior scores are positively correlated. This suggests that patients who display the behavioral pattern in Panel A also tend to show increased FA in the significant brain regions in Panel B. Panel D: Correlations between brain-behavior scores in the full sample (same as Panel C) and in held-out data using the cross-validation scheme described in methods.

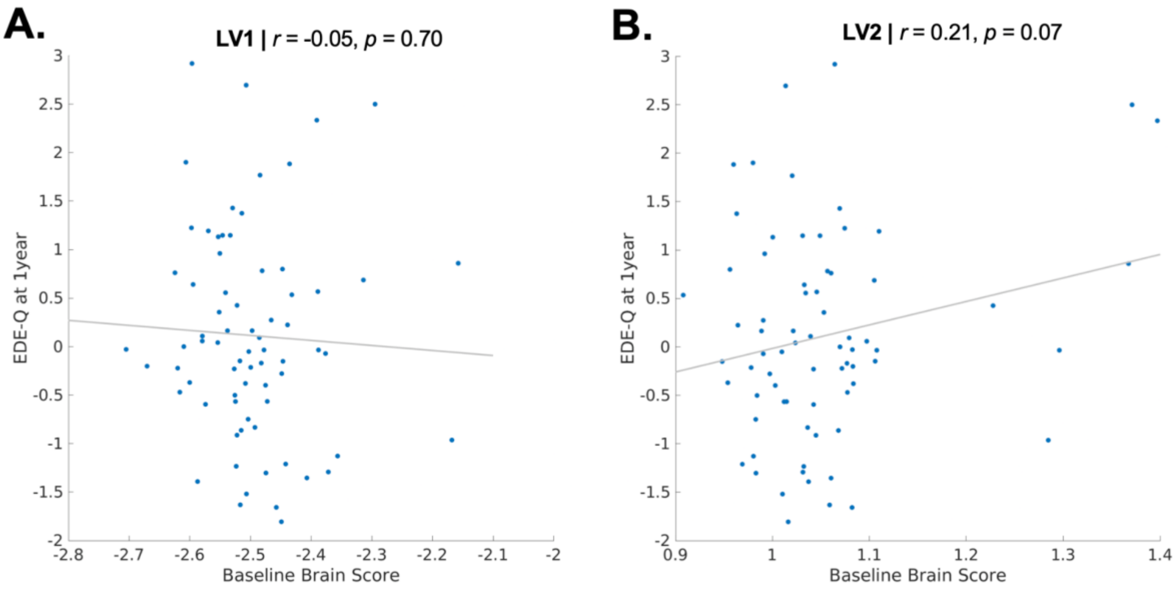


**Supplementary Figure 4.** Brain scores (Panel A: LV-1; Panel B: LV-2), derived from PLS analysis with FA brain measures, were not significantly associated with EDE-Q scores one year later, after adjusting for baseline EDE-Q scores.

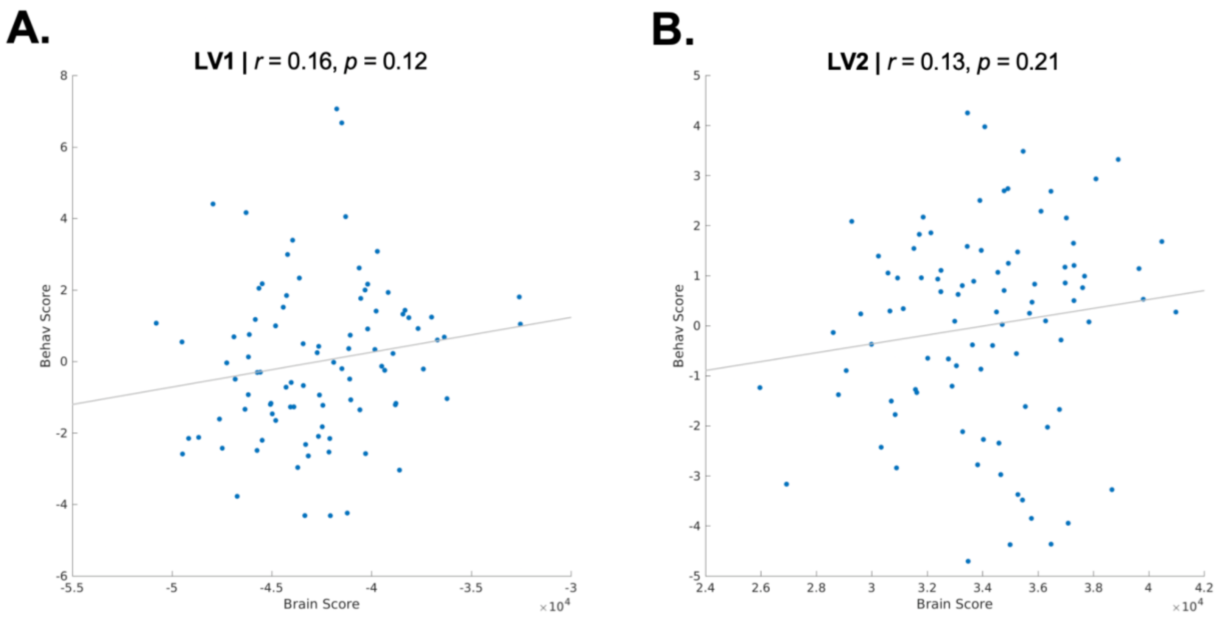


**Supplementary Figure 5.** Brain-behavior correlations for significant LVs (Panel A: LV-1; Panel B: LV-2) with cortical thickness/subcortical volume data used as input for the brain matrix for PLS. The top two LVs derived from these structural MRI features had significant permuted *p*-values, but did not show significant brain-behavior score associations. Further, these LVs did not survive permutation testing (LV1: out-of-sample *r*=-0.020, permuted *p*=0.53; LV2: out-of-sample *r*=-0.0051, permuted *p*=0.53). Thus, these results were not further interpreted in the main manuscript.
